## Supplementary Figure S1 for "Ascertaining the initiation of epidemic resurgences: an application to the COVID-19 second surges in Europe and the Northeast United States"

**Supplementary Figure S1. Location-specific characterization of the temporal evolution of the COVID-19 outbreak for the states in the Northeast US and countries in Europe's Schengen Area.**

(The 13-page long figure is appended after the caption)

The data for each location and for the region without the location is shown in a column of 4 panels. The 1st and 3rd panels from the top show the temporal evolution of the reproduction number (blue line) with the shaded blue region indicating the 95% confidence intervals (CI). The dotted lines highlight the span of the second wave and its confidence interval over the reproduction number data. The 2nd and 4th panels from the top show the temporal evolution of the infectious population with the shaded blue region indicating the 95% CI. The 1st and 2nd panels correspond to the location and the 3rd and 4th, to the region without the location. Reproduction numbers were computed from growth rates considering a gamma-distributed generation interval with a mean of 6.5 days and a standard deviation of 4.2 days. The trajectories of the infectious populations, the growth rates, and the 95%

confidence intervals (CI) for each location were downloaded on April 21, 2021, from https://github.com/Covid19Dynamics/trajectories.

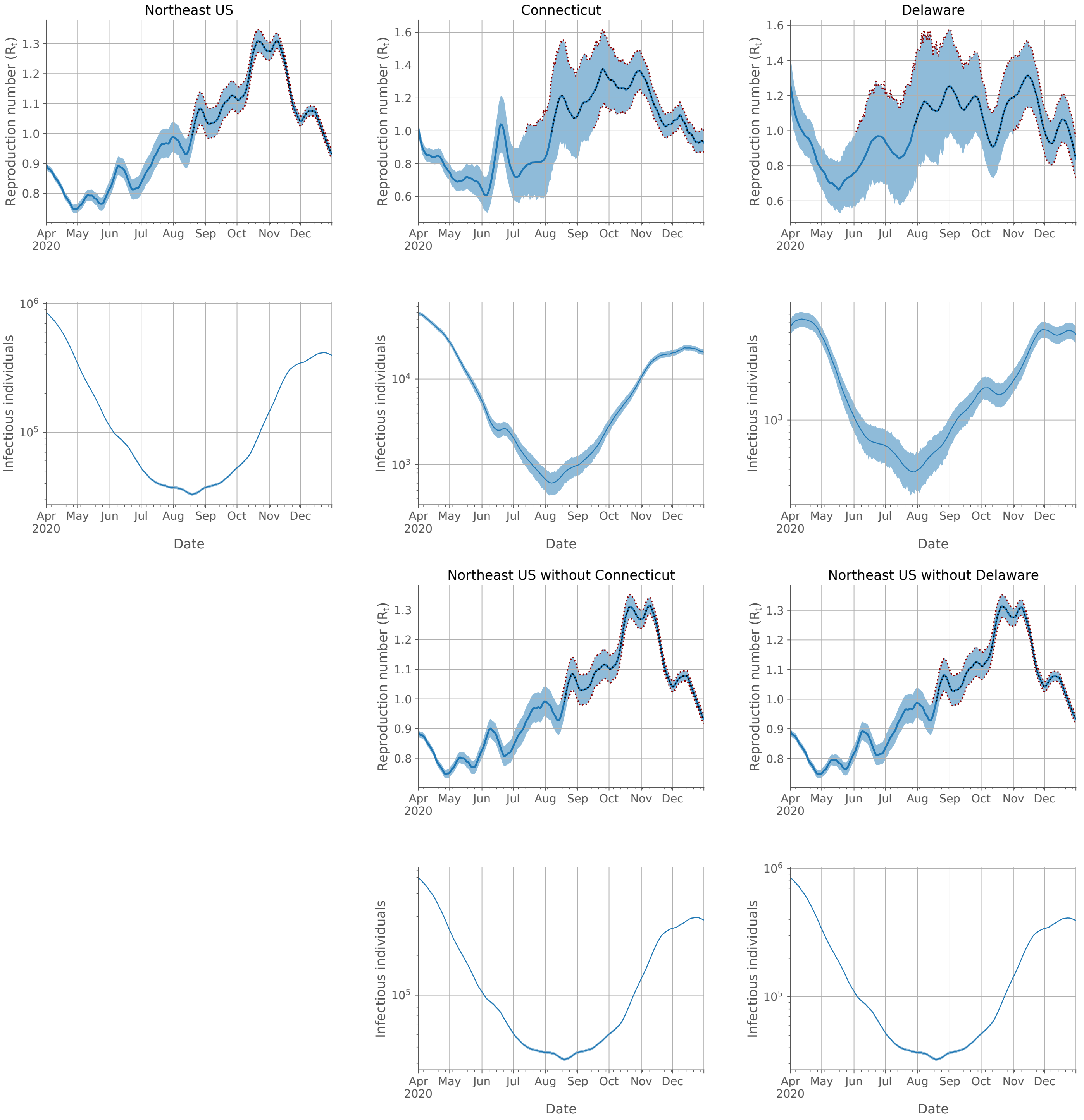


Supplementary Figure S1-1

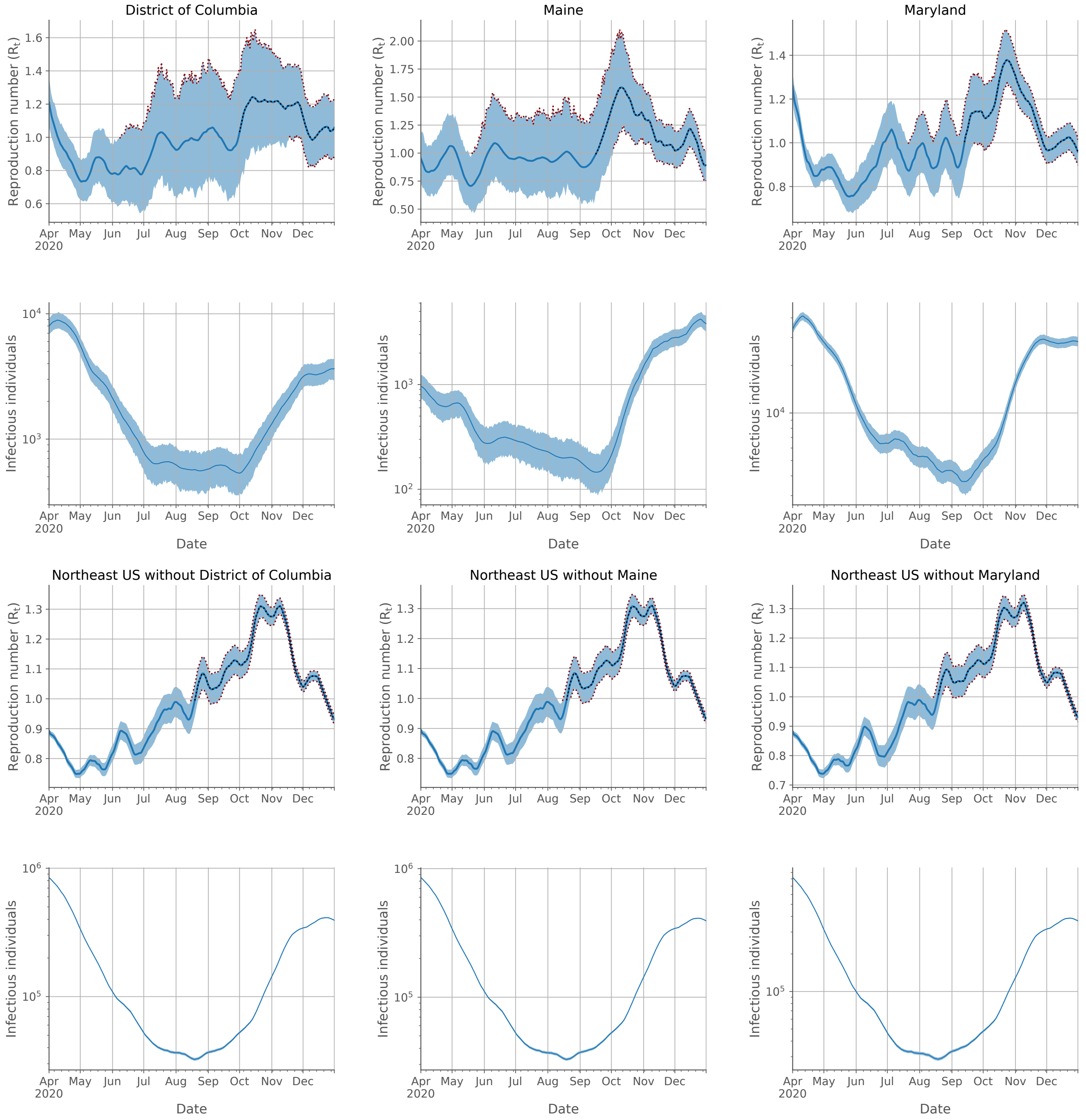


Supplementary Figure S1-2

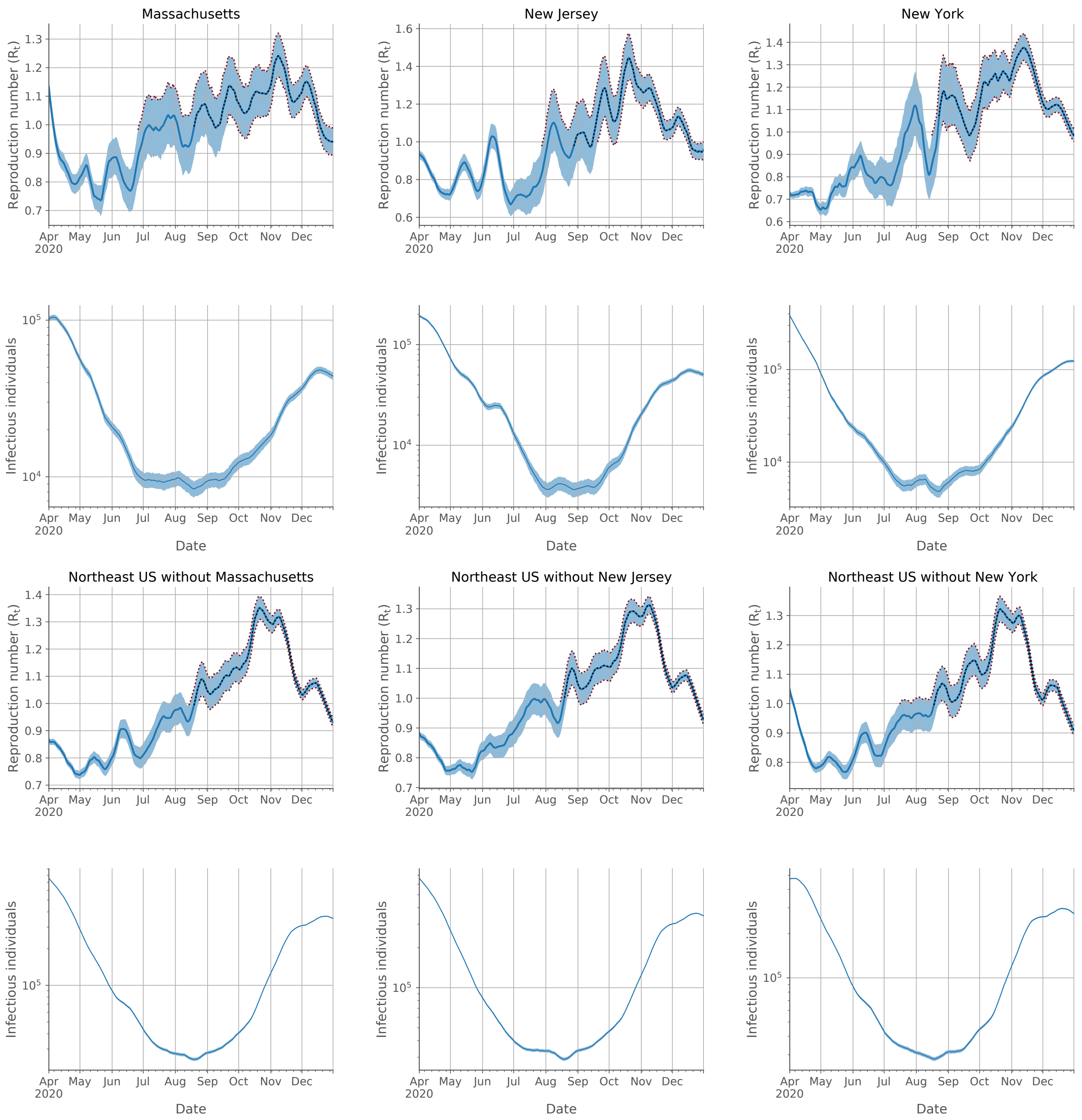


Supplementary Figure S1-3

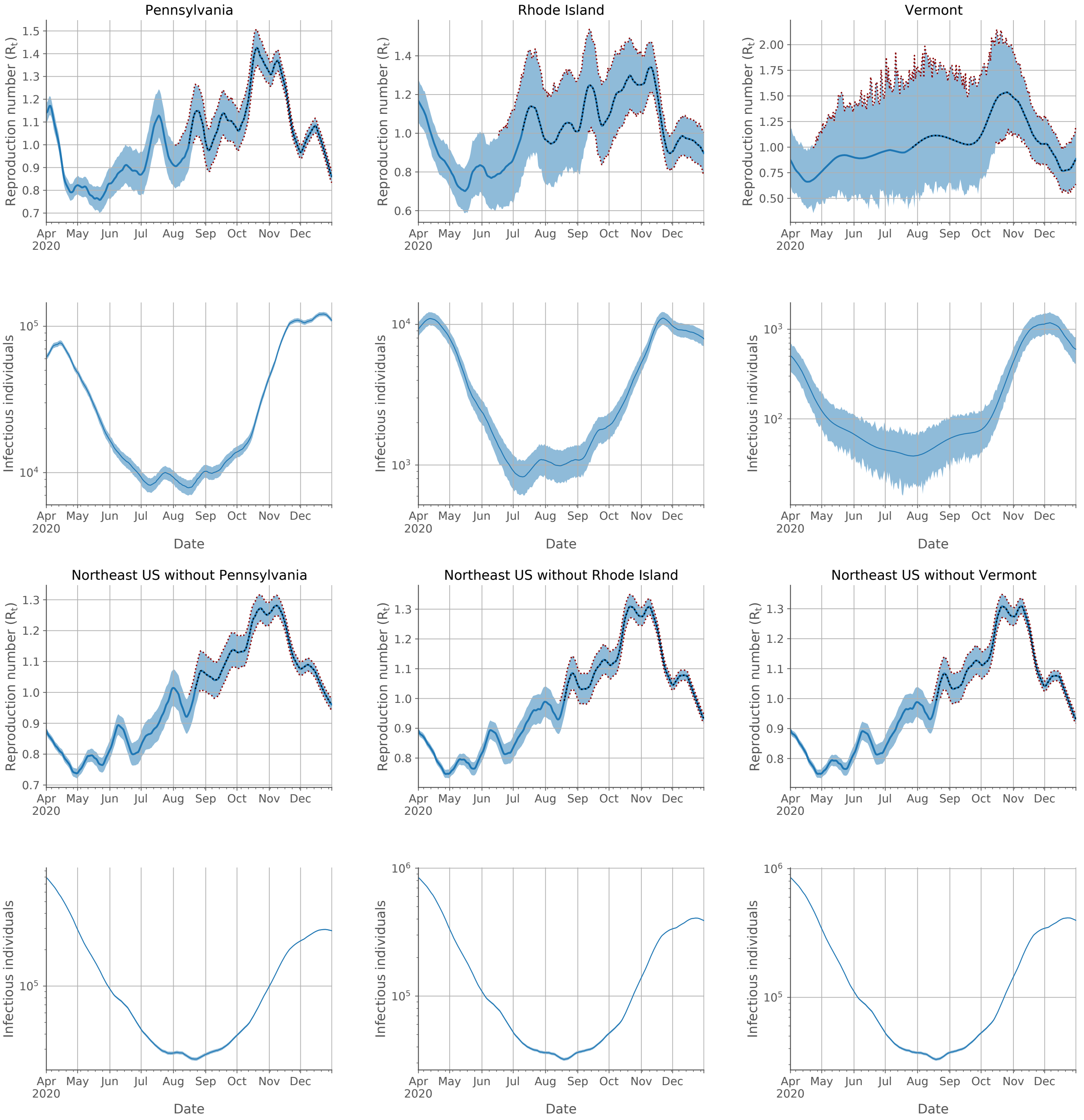


Supplementary Figure S1-4

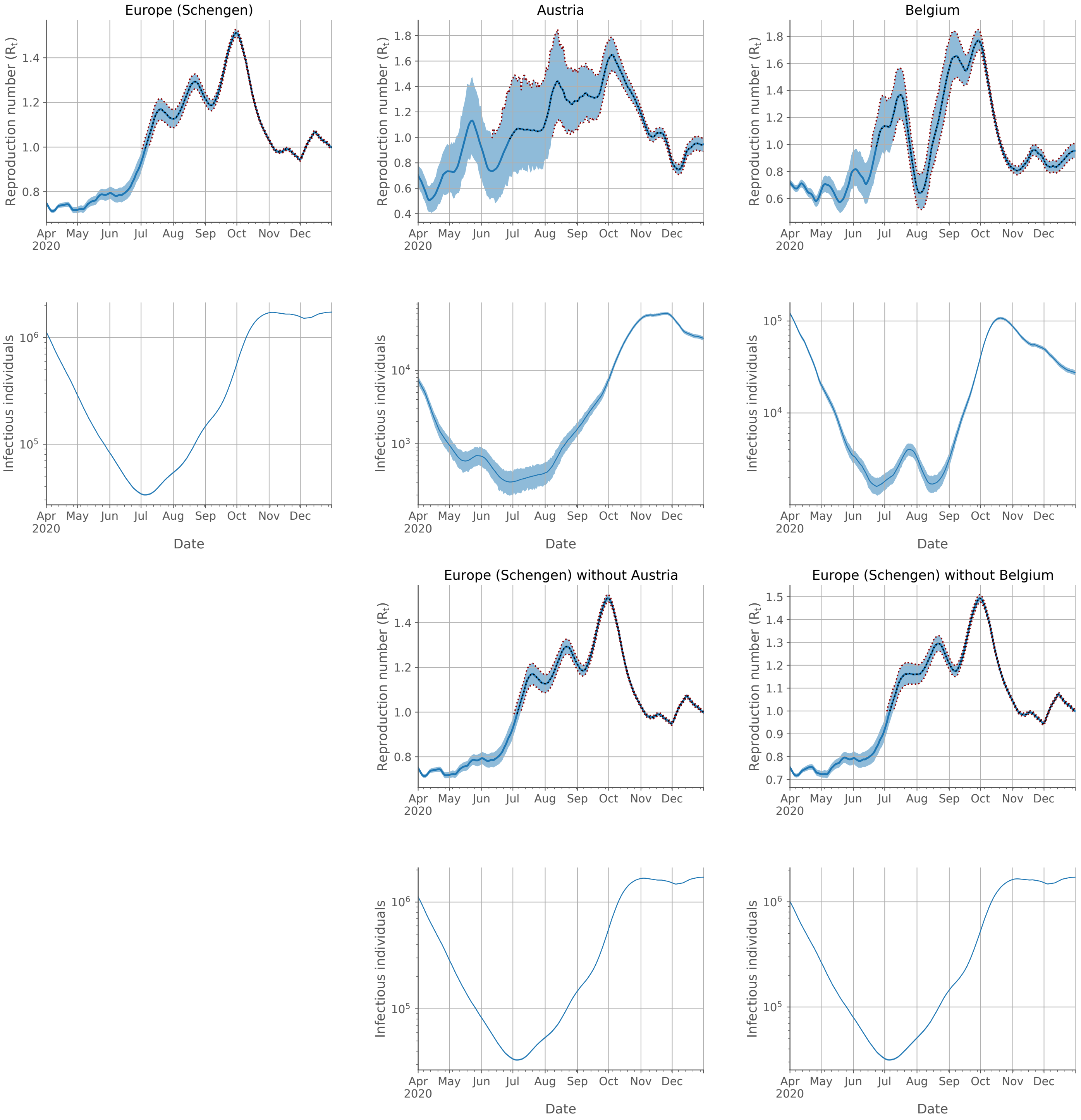


Supplementary Figure S1-5

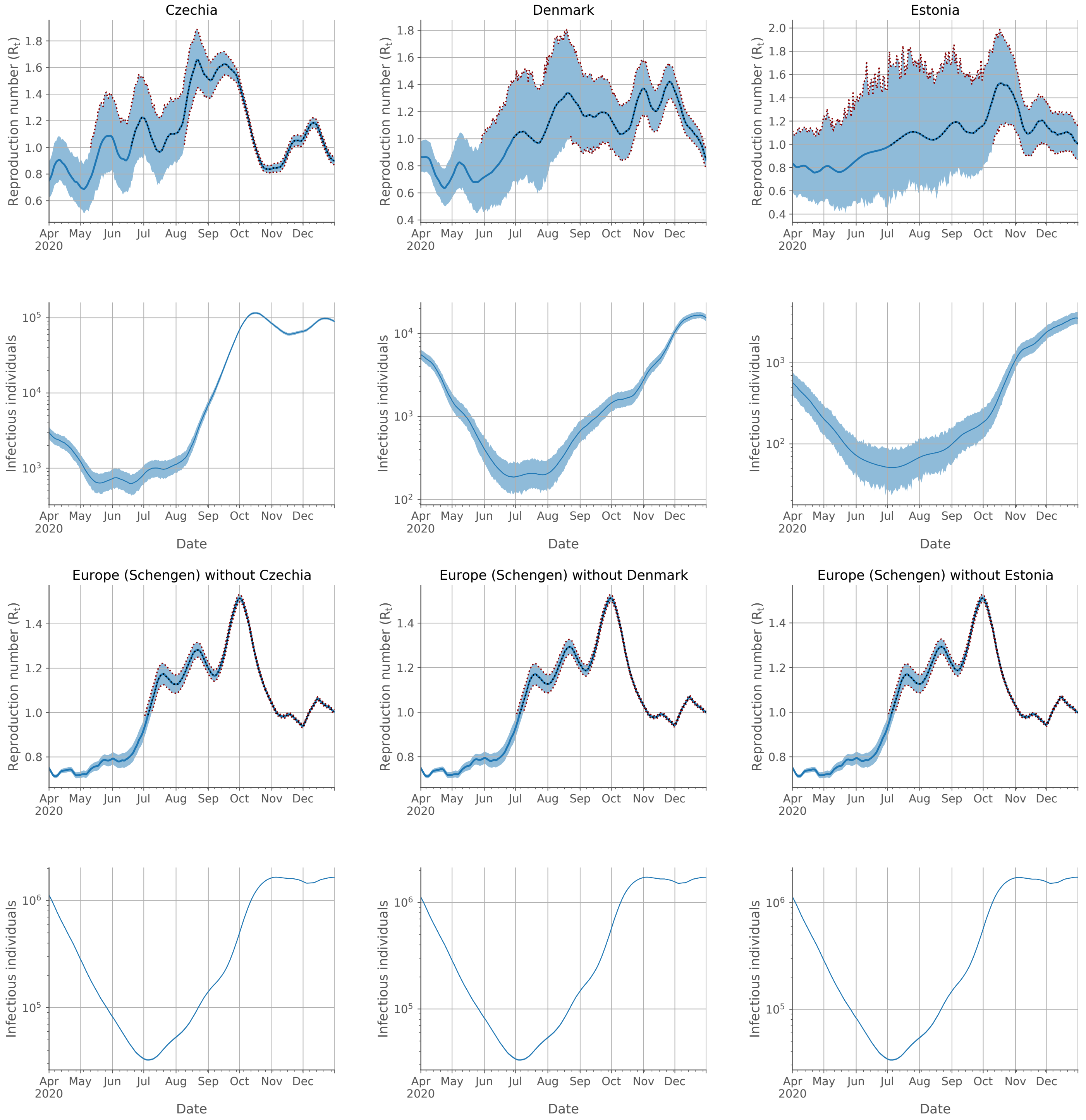


Supplementary Figure S1-6

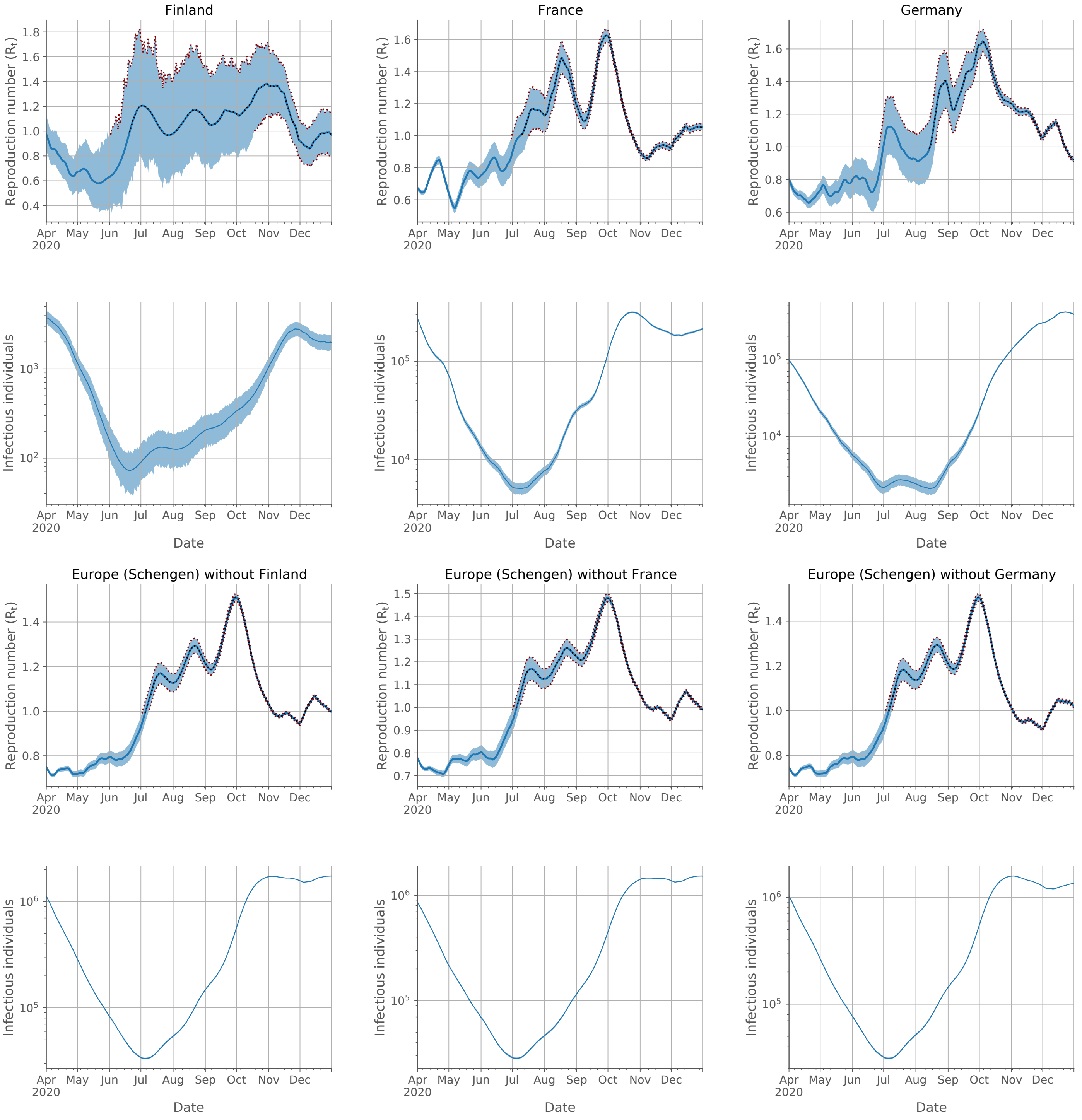


Supplementary Figure S1-7

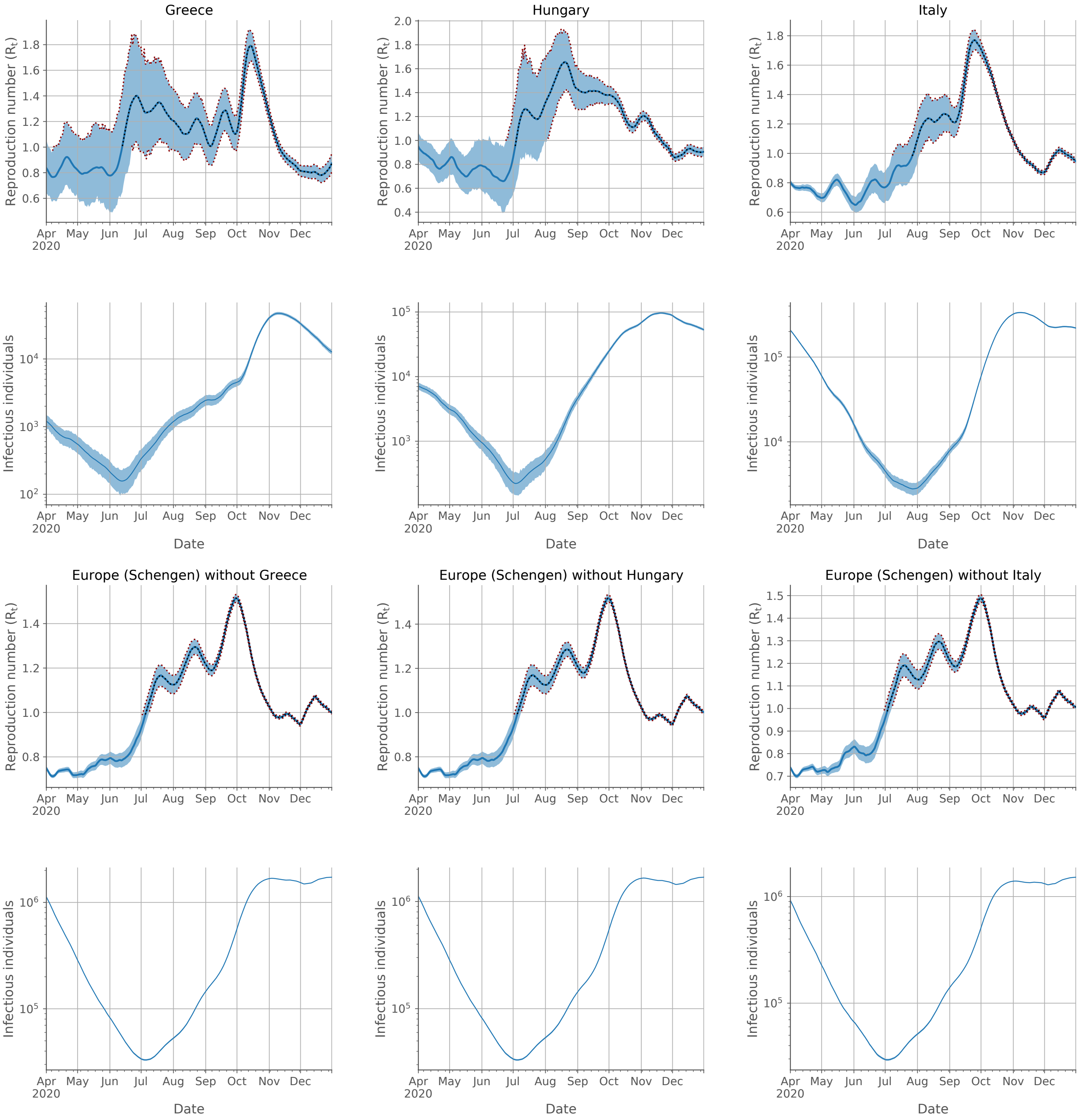


Supplementary Figure S1-8

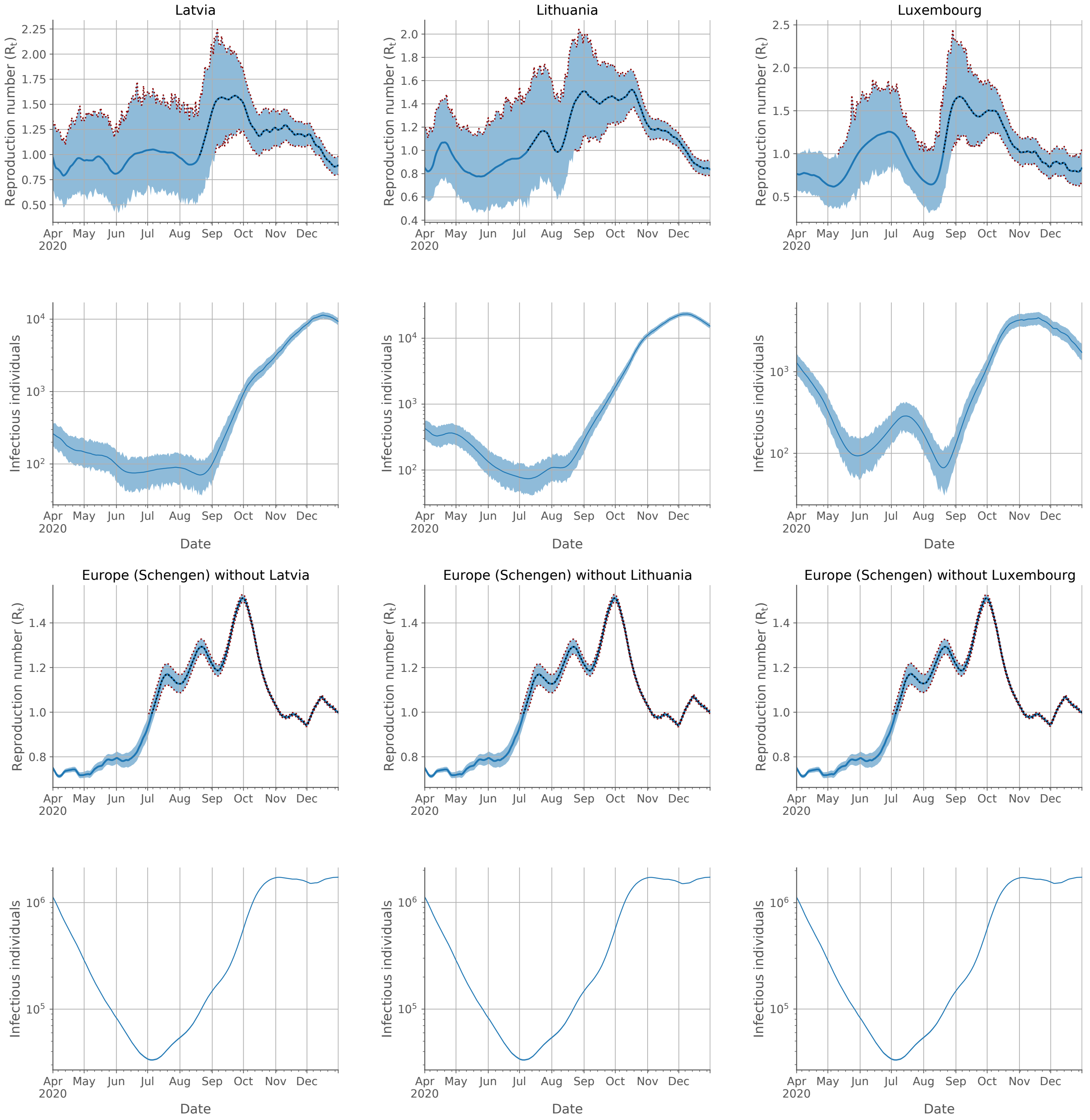


Supplementary Figure S1-9

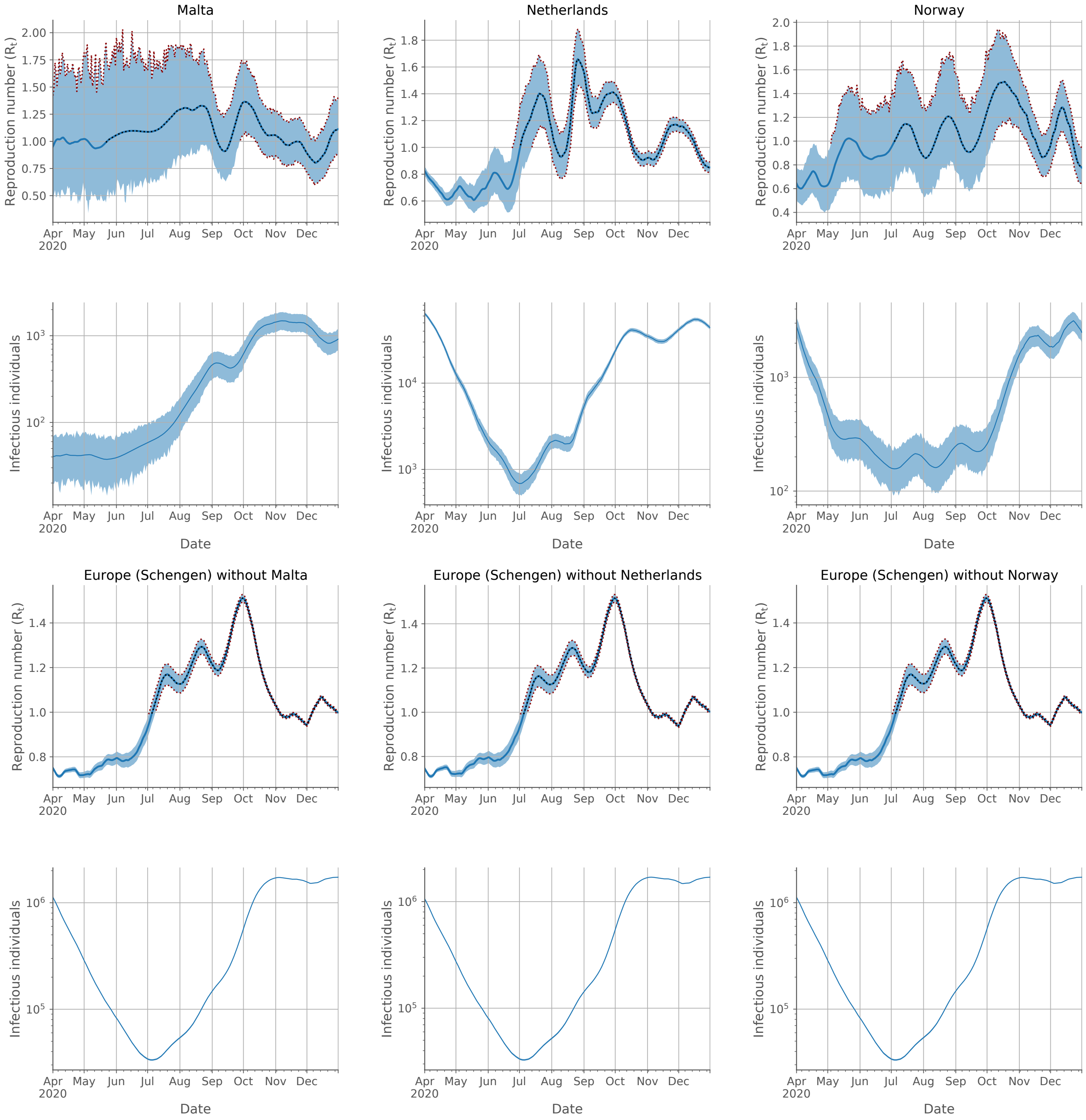


Supplementary Figure S1-10

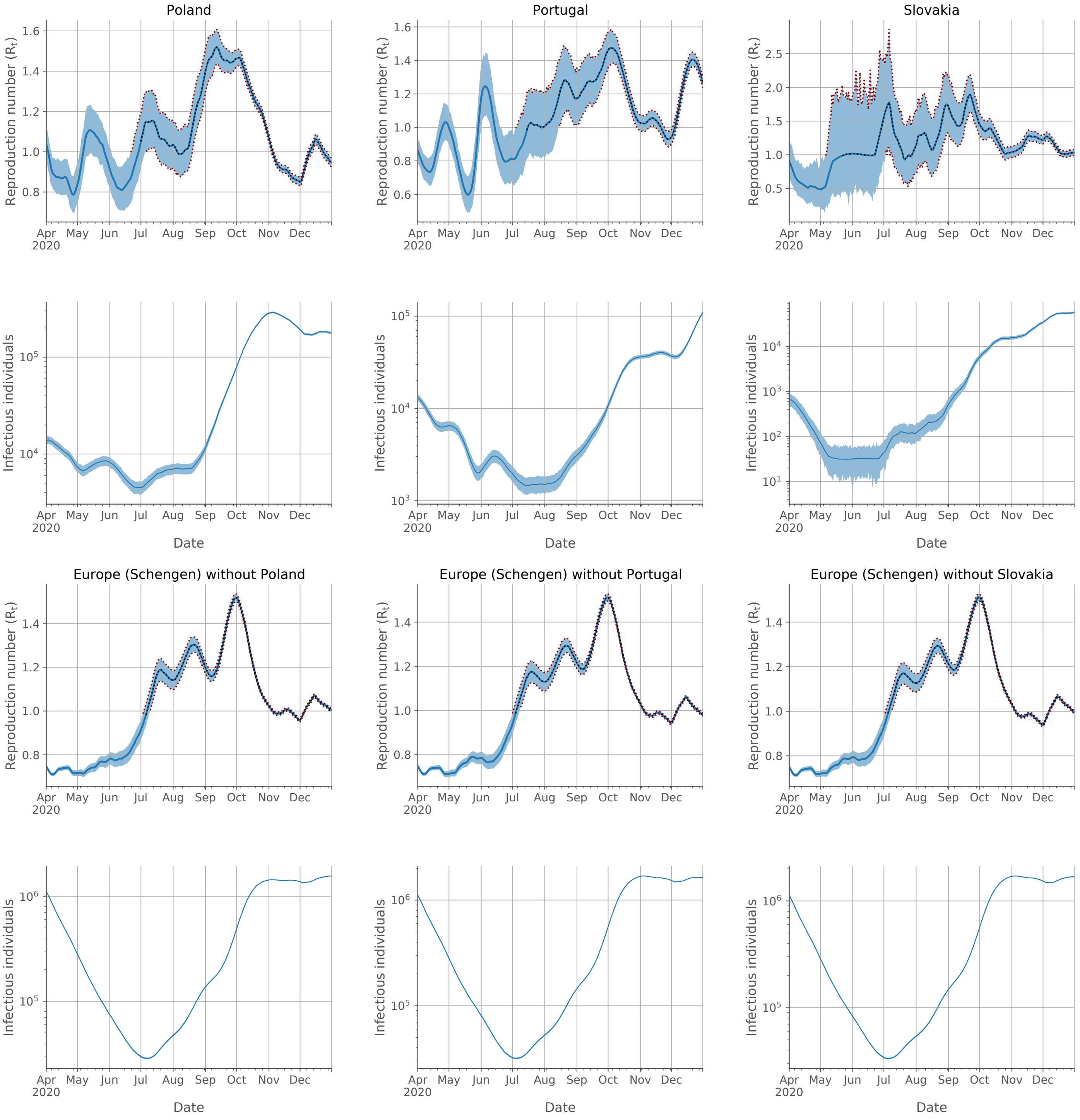


Supplementary Figure S1-11

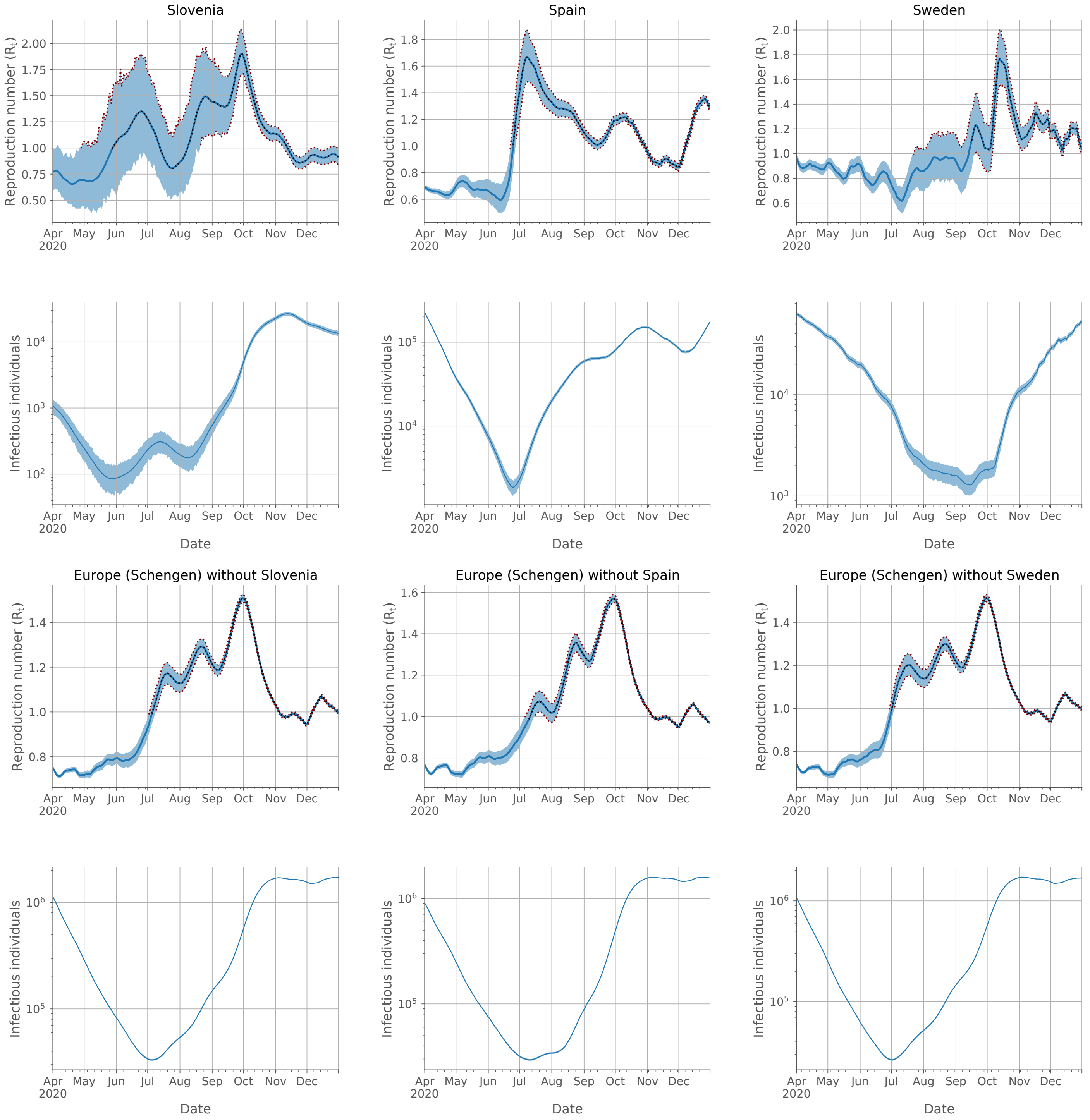


Supplementary Figure S1-12

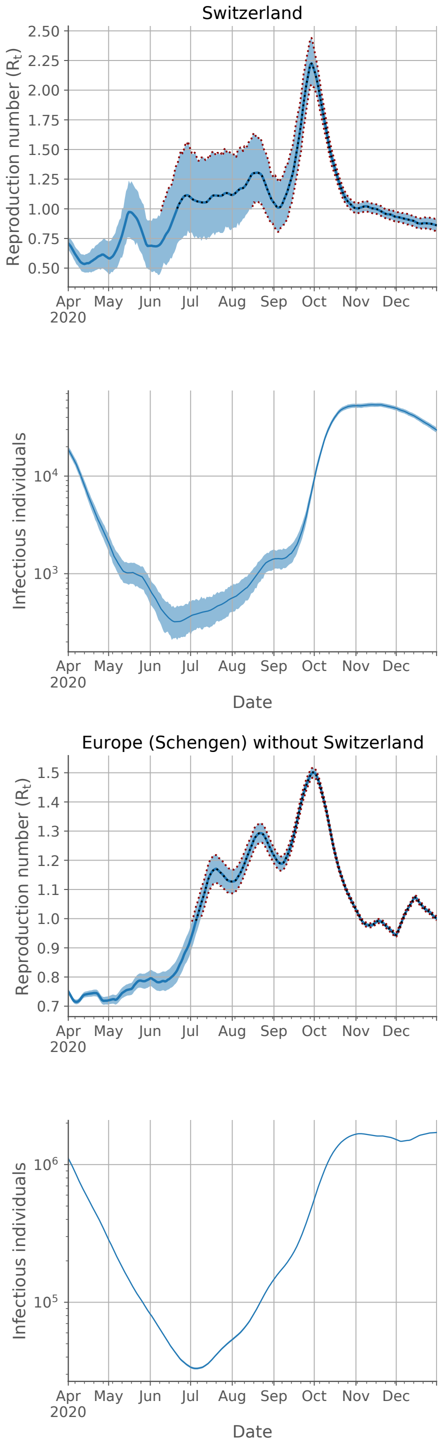


Supplementary Figure S1-13
